# Specific HLA Class I and II alleles are associated with a higher risk for tumor formation in Neurofibromatosis type 1

**DOI:** 10.64898/2026.05.04.26352173

**Authors:** Jonathan H. Sussman, Stephanie N. Brosius, Bernat Gel, Peiyao Li, Alvin Farrel, Jo Lynne Rokita, Edu Serra, Kai Tan, Michael J. Fisher, John M. Maris, Thomas De Raedt

## Abstract

Neurofibromatosis type 1 (NF1) is a common autosomal dominant genetic tumor predisposition syndrome.^1^ NF1 patients display remarkable phenotypic variability, even within families carrying the same *NF1* mutation.^2^ With few exceptions, the identification of specific genotype-phenotype correlations has remained elusive.^3–6^ We utilized RNA-seq data and direct DNA sequencing to determine HLA genotypes for individuals with NF1-associated high-grade glioma (HGG, n=25), low-grade glioma (LGG, n=79), and malignant peripheral nerve sheath tumors (MPNST, n=105). Odds ratios (OR), binomial p-values and false discovery values were calculated by comparing observed carrier frequencies against expected frequencies derived from ethnicity-matched population data. We find that specific HLA class I and II alleles are associated with different NF1 tumor types. For example, HLA-B*40:02 is significantly associated with NF1-MPNST (OR=3.71, p=0.001, Q=0.02), increasing the lifetime risks for MPNST from 10% to about 29%. The relative cancer risk for an individual in the general population carrying a risk allele can be high, however, that individual’s absolute risk for cancer typically remains very low. In contrast, individuals that carry a risk allele and are also burdened with a tumor predisposition syndrome will have a substantially higher absolute risk to develop a tumor, simply because they start at a higher baseline susceptibility for tumors. The identification of HLA-risk alleles for NF1 tumor development is therefore important, as it will allow for a risk-adapted screening or more aggressive treatment of individuals with a specific HLA haplotype. If confirmed, this study will thus improve clinical care and potential outcomes of individuals with NF1.

---

Neurofibromatosis type 1 is a complex disorder with both tumor and non-tumor manifestations. NF1 is one of the most frequent genetic disorders affecting about 1 in 3000 individuals. Although it is a multi-system disease, as a tumor predisposition syndrome, patients are at particular risk to develop benign neurofibromas, low-grade glioma (LGG, including both optic pathway glioma and non-optic pathway glioma), malignant peripheral nerve sheath tumors (MPNST, 10-15.8% life-time risk)^7,8^ and high-grade glioma (HGG, 1-2% lifetime risk), which include glioblastoma (GBM) and high-grade astrocytoma with piloid features (HGAP).^1,9,10^ It is important to note that manifestations of NF1 are very heterogeneous, even within members of the same family.^2^

Individuals with NF1 have a pathogenic germline variant in, or deletion of, the *NF1* tumor suppressor, which is a negative regulator of the RAS pathway. Somatic loss of the wild-type copy of *NF1* can lead to tumor formation and forms the basis of many of the features present in NF1 patients. Few recurrent germline variants in *NF1* exist, making genotype-phenotype correlations challenging. Specific germline variations in *NF1* do not explain the full heterogeneity of clinical manifestations observed in NF1.^2,11^ One of the rare examples of genotype-phenotype correlation is the increased risk for dermal neurofibromas and MPNST in those individuals with an *NF1* microdeletion (5%).^12^ The 1.4 Mb NF1 microdeletion encompasses *NF1* and surrounding areas, including the tumor suppressor *SUZ12* which is often mutated in MPNST.^13,14^ Specific germline pathogenic variants in *NF1* have been associated with large superficial plexiform and spinal neurofibromas (missense mutations in AA 844-848, which occur in 0.8% of NF1 individuals)^4^; others are associated with a Noonan-like phenotype (p.Met1149, p.Arg1276 and p.Lys1423, which represent about 1.8% of unrelated NF1 individuals)^6^. The p.Arg1276 pathogenic missense variants are associated with spinal neurofibromas; p.MET1149 individuals have a mild phenotype.^6^ It is important to note that collectively these alterations represent a small fraction of the NF1 population and do not explain the intra-familial variation of clinical manifestations.

Human leukocyte antigen (HLA) class I molecules, which include HLA-A, HLA-B, exhibit remarkable genetic diversity across human populations. This diversity is driven by the need to recognize and present a broad range of peptides (antigens) from intracellular pathogens, such as viruses, or neoantigens from mutated genes to CD8 T cells.^15^ The 3 classic HLA class I genes are located at the same genomic region (Chromosome 6p22.1) and each has multiple alleles. These alleles encode a slightly different peptide-binding groove, allowing for a wide range of antigenic peptides to be presented.^15^ It is noteworthy that all cells continuously interact with the immune system by displaying processed peptides on HLA receptors. These peptides can be derived from viral or mutated proteins but are most frequently from normal proteins that are expressed in the cell. This HLA display system is used by the immune system to recognize cells that are infected or unhealthy.^15^ The diversity of alleles for each HLA gene enhances the flexibility and adaptability of the immune system, but also influences susceptibility to diseases, including infections, autoimmune disorders, and cancer.^16^ HLA class II molecules on the other hand are typically expressed in antigen presenting immune cells, and play an important role in immune cell activation, including CD4 T-cells. Intriguingly, it has become evident that over 50% of NF1-associated HGG tumor cells express these HLA-class II genes that are typically restricted to immune cells.^17^ Expression of HLA-class II genes on non-immune cells is generally thought to be an immune suppressive. We therefore investigated if we would identify alleles from HLA-DRB1, the highest expressed HLA-Class II gene, which are associated with or underrepresented in specific NF1 tumors. In this study, we identified associations between alleles of both HLA Class I and II genes and the occurrence of HGG, LGG, and MPNST in individuals with Neurofibromatosis type 1.

As part of an immunotherapy research program at the Children’s Hospital of Philadelphia (CHOP), we HLA typed individuals with NF1 who developed HGG by using RNA-seq data analyzed with the arcasHLA algorithm^18^ v0.6.0 (default parameters) or by direct DNA sequencing (Supplementary Table 1) and observed an overrepresentation of some HLA alleles. Intrigued by these observations, we expanded our NF1-HGG cohort and investigated new cohorts of NF1-LGG and NF1-MPNST. We used published NF1 RNAseq data^19–26^ to determine HLA-alleles of individuals with NF1-HGG (25 patients^23,26^; Supplementary Table 1), NF1-LGG (79 patients^23–26^; Supplementary Table 2) and NF1-MPNST (105 patients^19–22,26,27^, Supplementary Table 3). Because the HLA frequencies vary across different populations, we used the race/ethnicity of each patient (White, Black, Asian, Hispanic) to calculate the expected frequency of each allele in each cohort. For some patients the race/ethnicity has been reported (CBTN), but for other patients the genetic ancestry was determined using whole genome or whole exome sequencing (Somalier algorithm using default settings) when available. This allowed us to determine the genetic ancestry of 88% of patients with NF1-HGG patients, 72% of patients with NF1-LGG patients and 67% of patients with NF1-MPNST (Supplementary Tables 1-3). In all cohorts we observed a strong bias towards White ethnicity. We combined the self-reported and determined genetic ancestry to infer the expected frequencies of all HLA alleles in the entire cohort of each tumor type. To investigate if identified HLA-alleles are also associated with the sporadic (non-tumor predisposition associated) counterparts of the NF1-associated tumors, we performed a similar exercise on patient data publicly available through the CBTN^26^ that sporadically developed pediatric HGG (pHGG, 204 patients, Supplementary Table 4) or pediatric LGG (pLGG, 218 patients; Supplementary Table 5) and included only those individuals in our study for whom reported race/ethnicity data is available. We realize that both the pHGG and pLGG cohorts are heterogeneous and contain different subtypes of glioma, which could affect our analysis. However, by comparing NF1-associated gliomas with sporadic gliomas, we are looking for HLA-alleles that are broadly underrepresented or associated with glioma formation. Of note, because we analyzed sequencing data derived tumor material, we cannot exclude that loss of heterozygosity (LOH) of Chromosome 6 (where the HLA genes are located) reduced the number of alleles identified in a patient. This could result in a slight undercounting of carriers of specific HLA alleles, which could lead to false positive identification of underrepresented alleles but simultaneously undercounting of alleles associated with tumor formation. MPNST and HGG, which are more prone to copy number variations (CNV) and LOH, have the highest risk of introducing this bias. LOH events are rare in NF1-associated LGG, reducing our concern for biases in that cohort. Moreover, only MPNST and not NF1-HGG nor NF1-LGG showed loss of Hardy-Weinberg equilibrium (Supplementary Table 6) of the HLA gene loci, suggesting some LOH took place in some MPNST tumors. Crucially, nearly all samples showed heterozygous HLA alleles across HLA-A, -B and/or C (24/25 NF1-HGG, 94/105 MPNST and 78/79 NF1-LGG), reducing our concern of an LOH bias in our analysis.

We determined the observed and expected percentage of patients that carry a specific HLA allele, calculated the odds ratio (OR) with confidence interval (CI), and binomial p-value (one sided). We also calculated the false discovery (Q) value for those alleles where at least 4 cases were observed or expected (NF1-HGG and NF1-LGG) or alleles with an observed or expected carrier frequency of at least 5% (MPNST, sporadic pediatric HGG, sporadic pediatric LGG). HLA-alleles with a false discovery rate lower than 25%, were considered significant. Alleles with a Q-value >0.25 but with a p-value <0.07 were considered borderline significant if the same allele is significant in any of the other cohorts. (Tables 1-5, Supplementary Table 7)

**Table 1.** HLA-I and II associations in NF1-HGG.

| NF1-HGG | A02:01 | A11 | B07:02 | B13:02 | B38:01 | B40:02 | C03:03 | C07:02 | C14:02 | DRB1-01:01 | DRB1-04:03 | DRB1-12:01 | DRB1-15:01 |
| --- | --- | --- | --- | --- | --- | --- | --- | --- | --- | --- | --- | --- | --- |
| cases (out of 25) | 7 | 7 | 5 | 4 | 0 | 1 | 6 | 5 | 1 | 3 | 0 | 0 | 10 |
| Patient carrier % | 26.9 | 26.9 | 19.2 | 15.4 | 0.0 | 3.8 | 23.1 | 19.2 | 3.8 | 12.5 | 0.0 | 0.0 | 41.7 |
| Expected carrier % | 44.1 | 12.0 | 21.6 | 4.9 | 2.1 | 3.0 | 9.4 | 24.8 | 3.0 | 15.9 | 1.5 | 3.3 | 24.7 |
| Odds Ratio | 0.47 | 2.70 | 0.87 | 3.52 | 0.00 | 1.29 | 2.88 | 0.72 | 1.30 | 0.76 | 0.00 | 0.00 | 2.18 |
| CI lower | 0.15 | 0.62 | 0.22 | 0.44 | NC | 0.06 | 0.58 | 0.19 | 0.06 | 0.15 | NC | NC | 0.64 |
| CI upper | 1.49 | 11.72 | 3.34 | 27.99 | NC | 26.05 | 14.27 | 2.70 | 26.60 | 3.87 | NC | NC | 7.48 |
| p-value | 0.056 | 0.030 | 0.497 | 0.037 | 0.569 | 0.549 | 0.031 | 0.344 | 0.545 | 0.457 | 0.695 | 0.452 | 0.051 |
| Q-value | 0.212 | 0.212 | 0.551 | 0.212 | NC | NC | 0.212 | 0.503 | NC | NC | NC | NC | 0.212 |
| Association | NEG | POS | NS | POS | NS | NS | POS | NS | NS | NS | NS | NS | POS |

CI: Confidence Interval, p-value: binomial p-value, Q-value: false discovery NC: not calculated, POS: positive association, NEG: underrepresentation, NS: not significant. HLA- DRB1 data was available for 24 samples. Green: underrepresentation, Orange: overrepresentation

**Table 2.** HLA-I and II associations in NF1-LGG.

| NF1-LGG | A02:01 | A11 | B07:02 | B13:02 | B38:01 | B40:02 | C03:03 | C07:02 | C14:02 | DRB1-01:01 | DRB1-04:03 | DRB1-12:01 | DRB1-15:01 |
| --- | --- | --- | --- | --- | --- | --- | --- | --- | --- | --- | --- | --- | --- |
| cases (out of 79) | 31 | 7 | 9 | 2 | 6 | 1 | 9 | 10 | 6 | 10 | 4 | 6 | 11 |
| Patient carrier % | 39.2 | 8.9 | 11.4 | 2.5 | 7.6 | 1.3 | 11.4 | 12.7 | 7.6 | 14.1 | 5.6 | 8.5 | 15.5 |
| Expected carrier % | 44.6 | 11.8 | 21.4 | 4.9 | 2.3 | 3.5 | 9.5 | 25.0 | 2.9 | 15.8 | 1.6 | 3.0 | 24.6 |
| Odds Ratio | 0.80 | 0.72 | 0.47 | 0.51 | 3.50 | 0.36 | 1.23 | 0.43 | 2.76 | 0.87 | 3.65 | 2.94 | 0.56 |
| CI lower | 0.43 | 0.26 | 0.20 | 0.09 | 0.64 | 0.04 | 0.44 | 0.19 | 0.58 | 0.35 | 0.44 | 0.60 | 0.24 |
| CI upper | 1.51 | 2.04 | 1.14 | 2.88 | 19.03 | 3.62 | 3.42 | 1.00 | 13.10 | 2.20 | 30.05 | 14.45 | 1.30 |
| p-value | 0.198 | 0.269 | 0.016 | 0.253 | 0.010 | 0.238 | 0.332 | 0.005 | 0.027 | 0.419 | 0.028 | 0.021 | 0.044 |
| Q-value | 0.459 | 0.459 | 0.245 | NC | 0.245 | NC | 0.503 | 0.245 | 0.245 | 0.514 | 0.245 | 0.245 | 0.294 |
| Association | NS | NS | NEG | NS | POS | NS | NS | NEG | POS | NS | POS | POS | NS |

CI: Confidence Interval, p-value: binomial p-value, Q-value: false discovery NC: not calculated, POS: positive association, NEG: underrepresentation, NS: not significant. HLA- DRB1 data was available for 71 samples. Green: underrepresentation, Orange: overrepresentation

**Table 3.** HLA-I and II associations in NF1-MPNST.

| MPNST | A02:01 | A11 | B07:02 | B13:02 | B38:01 | B40:02 | C03:03 | C07:02 | C14:02 | DRB1-01:01 | DRB1-04:03 | DRB1-12:01 | DRB1-15:01 |
| --- | --- | --- | --- | --- | --- | --- | --- | --- | --- | --- | --- | --- | --- |
| cases (out of 105) | 35 | 11 | 15 | 4 | 4 | 10 | 7 | 18 | 3 | 4 | 2 | 1 | 31 |
| Patient carrier % | 33.3 | 10.5 | 14.3 | 3.8 | 3.8 | 9.5 | 6.7 | 17.1 | 2.9 | 4.2 | 2.1 | 1.0 | 32.3 |
| Expected carrier % | 42.1 | 12.5 | 20.8 | 4.7 | 2.2 | 2.8 | 9.1 | 24.4 | 3.3 | 15.1 | 1.6 | 3.7 | 23.6 |
| Odds Ratio | 0.69 | 0.82 | 0.64 | 0.80 | 1.74 | 3.71 | 0.71 | 0.64 | 0.87 | 0.25 | 1.30 | 0.28 | 1.55 |
| CI lower | 0.39 | 0.35 | 0.31 | 0.21 | 0.34 | 0.97 | 0.26 | 0.33 | 0.18 | 0.08 | 0.16 | 0.03 | 0.82 |
| CI upper | 1.21 | 1.92 | 1.31 | 3.06 | 8.92 | 14.13 | 1.97 | 1.26 | 4.21 | 0.77 | 10.78 | 2.61 | 2.93 |
| p-value | 0.042 | 0.327 | 0.060 | 0.441 | 0.208 | 0.001 | 0.250 | 0.050 | 0.552 | 0.001 | 0.460 | 0.130 | 0.032 |
| Q-value | 0.299 | 0.441 | 0.299 | NC | NC | 0.017 | 0.410 | 0.299 | NC | 0.017 | NC | NC | 0.299 |
| Association | NS | NS | NS | NS | NS | POS | NS | NS | NS | NEG | NS | NS | NS |

CI: Confidence Interval, p-value: binomial p-value, Q-value: false discovery NC: not calculated, POS: positive association, NEG: underrepresentation, NS: not significant. HLA- DRB1 data was available for 96 samples. Green: underrepresentation, Orange: overrepresentation

**Table 4.** HLA-I and II associations of alleles significant in NF1 cohorts in sporadic pediatric HGG.

| Sporadic pHGG | A02:01 | A11 | B07:02 | B13:02 | B38:01 | B40:02 | C03:03 | C07:02 | C14:02 | DRB1-01:01 | DRB1-04:03 | DRB1-12:01 | DRB1-15:01 |
| --- | --- | --- | --- | --- | --- | --- | --- | --- | --- | --- | --- | --- | --- |
| cases (out of 204) | 79 | 18 | 37 | 11 | 9 | 7 | 15 | 53 | 10 | 24 | 5 | 12 | 37 |
| Patient carrier % | 38.7 | 8.8 | 18.1 | 5.4 | 4.4 | 3.4 | 7.4 | 26.0 | 4.9 | 11.8 | 2.5 | 5.9 | 18.1 |
| Expected carrier % | 41.9 | 12.2 | 19.4 | 4.5 | 2.8 | 4.0 | 8.9 | 24.4 | 3.2 | 14.1 | 2.0 | 3.3 | 22.4 |
| Odds Ratio | 0.87 | 0.70 | 0.92 | 1.22 | 1.61 | 0.84 | 0.81 | 1.08 | 1.58 | 0.81 | 1.22 | 1.84 | 0.77 |
| CI lower | 0.59 | 0.37 | 0.56 | 0.50 | 0.55 | 0.30 | 0.40 | 0.69 | 0.58 | 0.45 | 0.33 | 0.70 | 0.47 |
| CI upper | 1.30 | 1.32 | 1.51 | 3.00 | 4.68 | 2.35 | 1.66 | 1.70 | 4.34 | 1.45 | 4.56 | 4.83 | 1.25 |
| p-value | 0.195 | 0.083 | 0.361 | 0.304 | 0.120 | 0.414 | 0.264 | 0.330 | 0.115 | 0.194 | 0.395 | 0.038 | 0.084 |
| Q-value | 0.470 | 0.337 | 0.470 | 0.470 | NC | NC | 0.470 | 0.337 | NC | 0.470 | NC | 0.337 | 0.337 |
| Association | NS | NS | NS | NS | NS | NS | NS | NS | NS | NS | NS | NS | NS |

CI: Confidence Interval, p-value: binomial p-value, Q-value: false discovery NC: not calculated, POS: positive association, NEG: underrepresentation, NS: not significant. Green: underrepresentation, Orange: overrepresentation

**Table 5.** HLA-I and II associations of alleles significant in NF1 cohorts in sporadic pediatric LGG.

| Sporadic pLGG | A02:01 | A11 | B07:02 | B13:02 | B38:01 | B40:02 | C03:03 | C07:02 | C14:02 | DRB1-01:01 | DRB1-04:03 | DRB1-12:01 | DRB1-15:01 |
| --- | --- | --- | --- | --- | --- | --- | --- | --- | --- | --- | --- | --- | --- |
| cases (out of 218) | 90 | 22 | 37 | 21 | 7 | 11 | 16 | 48 | 9 | 27 | 10 | 7 | 37 |
| Patient carrier % | 41.3 | 10.1 | 17.0 | 9.6 | 3.2 | 5.0 | 7.3 | 22.0 | 4.1 | 12.4 | 4.6 | 3.2 | 17.0 |
| Expected carrier % | 43.5 | 11.0 | 21.0 | 4.7 | 2.3 | 3.3 | 9.0 | 24.3 | 2.9 | 15.2 | 1.5 | 3.3 | 23.5 |
| Odds Ratio | 0.91 | 0.91 | 0.77 | 2.18 | 1.42 | 1.55 | 0.80 | 0.88 | 1.46 | 0.79 | 3.17 | 0.96 | 0.67 |
| CI lower | 0.62 | 0.49 | 0.48 | 1.01 | 0.44 | 0.60 | 0.40 | 0.56 | 0.52 | 0.46 | 0.89 | 0.34 | 0.42 |
| CI upper | 1.33 | 1.67 | 1.24 | 4.73 | 4.57 | 4.05 | 1.59 | 1.38 | 4.14 | 1.36 | 11.23 | 2.77 | 1.07 |
| p-value | 0.276 | 0.382 | 0.082 | 0.001 | 0.231 | 0.110 | 0.233 | 0.245 | 0.175 | 0.144 | 0.002 | 0.561 | 0.012 |
| Q-value | 0.387 | 0.445 | 0.237 | 0.036 | NC | 0.258 | 0.375 | 0.375 | NC | 0.307 | NC | NC | 0.089 |
| Association | NS | NS | NEG | POS | NS | NS | NS | NS | NS | NS | POS | NS | NEG |

CI: Confidence Interval, p-value: binomial p-value, Q-value: false discovery NC: not calculated, POS: positive association, NEG: underrepresentation, NS: not significant. Green: underrepresentation, Orange: overrepresentation

We found an overrepresentation (Q<0.25) of 3 HLA-class I alleles in our cohort of 25 NF1-HGG patients (HLA-A*11, HLA-B*13:02, and HLA-C*03:03, Table 1). None of these were significant in the sporadic pediatric HGG population (Table 4) but HLA-B*13:02 was associated with sporadic pediatric LGG (Table 5). Two noteworthy alleles are HLA-A*11 and HLA-B*13:02 with an OR of 2.70 and 3.52 respectively. Of the 7 patients carrying A*11 allele, 4 were under 18 years of age, which is noteworthy given how rare NF1-HGG are in the pediatric population. However, in our cohort of 25 patients, we do not observe a significant difference between the age of HGG diagnosis in patients that carry HLA-A*11 and those that do not (4/7 <18yo with A*11 versus 7/14 without A*11, p=0.39, Fisher Exact test). The large number of pediatric patients that carry HLA-A*11 is likely due to ascertainment bias, as many of the samples of NF1 patients in our study were obtained from the CBTN (15/25). Intriguingly, 3 of 7 patients carrying HLA-A*11 also carried HLA-B*13:02. It is possible that both alleles form one haplotype, however, we did not find strong evidence that A*11-B*13:02 is a common haplotype in most across different genetic ancestries. It was not listed as a common haplotype in studies of the Kinh Vietnamese or Caucasian populations; however, in the Guizhou province in China the A*11-B*13:02 haplotype frequency is 3.94%. In our NF1-HGG cohort, only one individual was of Asian descent. These results need to be validated in a new larger cohort of NF1-HGG patients ideally with both demographic and outcome data. One HLA-class I allele was underrepresented (Q<0.25) in our NF1-HGG cohort (HLA-A*02:01, OR=0.47). Interestingly, HLA-A*02:01 also had a significant p-value in MPNST (p=0.042, Q=0.299).

Table 2 lists the HLA alleles that are associated (B*38:01, OR=3.50, p=0.010, Q=0.245) or underrepresented (B*07:02, OR=0.47, p=0.016, Q=0.245 and C*07:02, OR=0.43, p=0.005, Q=0.245) in NF1 patients with LGG. Of note, HLA-C*07:02 also had a borderline significant p-value in our MPNST cohort even though the Q-value did not reach significance (OR=0.64, p=0.05, Q=0.299). Allele B*07:02 was also underrepresented in sporadic pediatric HGG (Table 5).

A single HLA-allele reached significance in MPNSTs; allele HLA-B*40:02 was carried in 10 of 105 NF1-MPNST (9.5%), while we expect only 2.8% of individuals to carry this allele (OR=2.76, CI=1.0-14.1; p=0.001, binomial distribution, Q=0.017) (Table 3). The allele frequency of HLA-B*40:02 is uniformly low across races (Caucasian 1.3%, African American 0.3%, Hispanic 4.9%, Asian 2.4%). This association between HLA-B*40:02 and NF1-MPNST means that carriers of HLA-B*40:02 have an estimated lifetime risk for MPNST of 29.2-41.0% (calculated using Bayes’ Theorem) which is substantially higher than the 10%-15.8% lifetime risk^7,8^ of the general NF1 population. Strikingly, this allele is especially frequent in NF1 individuals who developed an MPNST before the age of 18. Of the 9 pediatric patients in our NF1-MPNST cohort, 4 carry HLA-B*40:02, which is significantly higher than the number of pediatric MPNST patients among individuals with NF1 that do not carry HLA-B*40:02 (p=0.0027, Fisher exact test, Age of diagnosis data was available for 34/105 MPNST patients). The MPNST study included patients from several hospitals/institutes reducing the risk of ascertainment bias on age^19,21,22,27^.

We performed a similar analysis for HLA-class II gene DRB1, which is typically only expressed on professional antigen presenting immune cells, but in NF1 tumors is expressed on tumor cells. We identified 4 alleles that were significantly associated with or underrepresented in NF1-associated tumors (Table 1-3). Noteworthy is HLA-DRB1*15:01, which is associated with NF1-HGG (OR=2.18, CI=0.64-7.48, p=0.051, Q=0.404), and borderline significantly associated with MPNST (OR=1.55, CI=0.82-2.93, p=0.032, Q=0.299). Surprisingly, HLA-DRB1*15:01 is underrepresented in sporadic pediatric LGG (OR=0.67, p=0.012, Q=0.089) and close to being significantly underrepresented in NF1-LGG (OR=0.56, CI=0.24-1.30, p=0.044, Q=0.294). Interestingly, HLA-DRB1*01:01 is highly significantly underrepresented in NF1-MPNST patients (OR=0.25, CI=0.08-0.77, p<0.001, Q=0.009), suggesting this is a protective allele in MPNST. Finally, we found that 2 HLA-DRB1 alleles are associated with NF1-LGG (DRB1*04:03, OR=3.65, p=0.028, Q=0.245 and DRB1*12:01, OR=2.94, p=0.021, Q=0.245). DRB1*04:03 is also enriched in the sporadic pediatric LGG population (OR=3.17, CI=0.89-11.23, p=0.002).

We have identified several HLA-alleles that potentially alter the susceptibility of NF1 individuals to develop NF1-HGG, NF1-LGG or MPNST. However, it remains unknown why specific HLA alleles or haplotypes are associated with NF1 tumor formation. We consider three hypotheses: (1) Infection susceptibility: Specific alleles can predispose patients to viral or bacterial infections, which in turn can contribute to tumor formation. For example, associations between A*11 and chronic hepatitis C virus as well as A*11 and severity of COVID have been described.^28–30^ Additionally, specific HLA alleles enhance the risk for human papillomavirus persistence and initiation of cervical cancer^31^. However, as far as we are aware, no viral infections have been associated with tumor formation in NF1. (2) Ability of HLA-class I alleles to display tumor specific peptides: The primary function of HLA class I receptors is to present peptides to which the immune system can respond. Alleles associated with tumor formation could have a reduced ability to display specific tumor peptides that trigger the immune system. However, one would expect that the other HLA alleles would compensate for this deficiency, making this hypothesis of how HLA-alleles are associated with tumor formation less likely. In contrast, underrepresentation of HLA alleles in patients with cancer (i.e., protection against a tumor), can be explained by the efficient presentation/binding of a tumor-specific peptides/neoantigens in the grove of the HLA receptor to elicit a T-cell response. (3) Drivers of tumor formation closely linked to HLA-genes: A polymorphism or gene variant closely linked to the associated HLA haplotype could predispose to tumor formation in NF1. Several genes, other than HLA genes, are present in the MHC genomic locus where all HLA Type I genes reside. One of them, *POU5F1* (*OCT4*), is of interest as it is an important transcription factor with a role in cancer stem cells.^32^ *POU5F1* is located within 290kb from HLA-B and C, and 1.2Mb from HLA-A. This close physical location indicates they could be in linkage disequilibrium.

HLA-Class II receptors are typically only presented by antigen presenting immune cells; however, recent data shows that a substantial number of NF1-HGG tumor cells express the HLA-class II antigen presenting machinery.^17^ Similar observations were made in other cancers, although the percentage of tumor cells that display HLA-class II molecules is substantially lower than what was observed in NF1-HGG^33–35^. The specific mechanism of how HLA-class II expression on tumor cells contributes to tumor formation remains elusive, but it is suggested they play an immune suppressive role through stimulation of CD4 T-cells and stimulation of exhaustion of CD8 T-cells. It also remains unclear whether the presentation of specific antigens in HLA-class II molecules is important. Specific HLA-class II alleles can thus have a tumor promoting or protecting role through expression on immune cells or tumor cells.

It is known that HLA alleles are associated with or underrepresented in patients with cancer. In the general population the actual lifetime risk for cancer when you carry such an associated allele usually remains low, as the observed ORs are often in the 1-3 range and the baseline lifetime risk for a specific cancer itself is very low. However, in patients that have an elevated baseline risk to develop tumors, like individuals with NF1, this modest change in OR can dramatically affect outcomes. The association of HLA alleles with tumor formation in predisposition syndromes like NF1 can thus have a direct impact on patient care. Particularly for NF1-HGG and MPNST, the odds ratios of the observed associations suggest a dramatic increase in the likelihood of an NF1 patient developing malignancies. HLA-B*13:02 for example would increase the lifetime risk for HGG from 1-2% to up to 6.7%; while HLA-B*40:02 could increase the lifetime risk for MPNST up to 40%. The HLA data reported here warrant further research into HLA genotype-phenotype associations in Neurofibromatosis type 1. This report is a call to action for the community to consider HLA-typing of NF1 individuals and to validate these observations in a large HLA genotype-phenotype study, with the hope that one day HLA data may improve the clinical care of those with NF1.

## Data Availability

All HLA data is available in supplemental tables. Original/source data used in the paper is available is publicly available: d Angelo et al. 2019, Fisher et al. 2021, Kochat et al. 2021, GeM consortium (EGAD00001008608, https://doi.org/10.7303/syn23651229), Yan et al. 2022, Pan et al. 202125, CBTN (https://cbtn.org/pediatric-brain-tumor-atlas, https://pedcbioportal.org/study/summary?id=openpbta, https://cavatica.sbgenomics.com/u/cavatica/pbta-cbttc), Larsson et al. 2023. This study did not generate additional datasets or code.

https://pedcbioportal.org/study/summary?id=openpbta

https://cavatica.sbgenomics.com/u/cavatica/pbta-cbttc

## Declaration of interests

The authors declare no competing interests.

## Acknowledgements

We thank the patients and their families for donating data to the biorepositories used in this study.

## Funding

Gilbert Family Foundation, The Children’s Tumor Foundation, DHART SPORE, Alex’s Lemonade Stand Foundation, Jeff Gorden Pediatric Research Foundation, NIH K12.

## Author contributions

J.H.S., S.N.B., B.G., P.Y., E.S., A.F, K.T., M.J.F., J.M., T.D.R., performed analysis and contributed to data interpretation. JLR advised on methodology, reviewed and edited the manuscript, J.H.S., S.N.B and T.D.R. wrote the paper.

## Data and code availability

Original/source data used in the paper is available is publicly available: d’Angelo *et al*. 2019^23^, Fisher *et al*. 2021^24^, Kochat *et al*. 2021^19^, GeM consortium^27^ (EGAD00001008608, https://doi.org/10.7303/syn23651229), Yan *et al*. 2022^22^, Pan *et al*. 2021^25^, CBTN^26^ (https://cbtn.org/pediatric-brain-tumor-atlas, https://pedcbioportal.org/study/summary?id=openpbta, https://cavatica.sbgenomics.com/u/cavatica/pbta-cbttc), Larsson *et al*. 2023^21^. This study did not generate additional datasets or code.

## Notes

### Competing Interest Statement

The authors have declared no competing interest.

### Funding Statement

Gilbert Family Foundation, The Childrens Tumor Foundation, DHART SPORE, Alexs Lemonade Stand Foundation, Jeff Gorden Pediatric Research Foundation, NIH K12

### Author Declarations

Original/source data used in the paper is available is publicly available: d Angelo et al. 2019, Fisher et al. 2021, Kochat et al. 2021, GeM consortium (EGAD00001008608, https://doi.org/10.7303/syn23651229), Yan et al. 2022, Pan et al. 202125, CBTN (https://cbtn.org/pediatric-brain-tumor-atlas, https://pedcbioportal.org/study/summary?id=openpbta, https://cavatica.sbgenomics.com/u/cavatica/pbta-cbttc), Larsson et al. 2023. This study did not generate additional datasets or code.

## References

1. Gutmann, D.H., Ferner, R.E., Listernick, R.H., Korf, B.R., Wolters, P.L., and Johnson, K.J. (2017). Neurofibromatosis type 1. Nat Rev Dis Primers 3, 17004. 10.1038/nrdp.2017.4.

2. Sabbagh, A., Pasmant, E., Laurendeau, I., Parfait, B., Barbarot, S., Guillot, B., Combemale, P., Ferkal, S., Vidaud, M., Aubourg, P., et al. (2009). Unravelling the genetic basis of variable clinical expression in neurofibromatosis 1. Hum Mol Genet 18, 2768–2778. 10.1093/hmg/ddp212.

3. Ottenhoff, M.J., Rietman, A.B., Mous, S.E., Plasschaert, E., Gawehns, D., Brems, H., Oostenbrink, R., Team, E.-N., van Minkelen, R., Nellist, M., et al. (2020). Examination of the genetic factors underlying the cognitive variability associated with neurofibromatosis type 1. Genet Med 22, 889–897. 10.1038/s41436-020-0752-2.

4. Koczkowska, M., Chen, Y., Callens, T., Gomes, A., Sharp, A., Johnson, S., Hsiao, M.C., Chen, Z., Balasubramanian, M., Barnett, C.P., et al. (2018). Genotype-Phenotype Correlation in NF1: Evidence for a More Severe Phenotype Associated with Missense Mutations Affecting NF1 Codons 844-848. Am J Hum Genet 102, 69–87. 10.1016/j.ajhg.2017.12.001.

5. Napolitano, F., Dell’Aquila, M., Terracciano, C., Franzese, G., Gentile, M.T., Piluso, G., Santoro, C., Colavito, D., Patane, A., De Blasiis, P., et al. (2022). Genotype-Phenotype Correlations in Neurofibromatosis Type 1: Identification of Novel and Recurrent NF1 Gene Variants and Correlations with Neurocognitive Phenotype. Genes (Basel) 13. 10.3390/genes13071130.

6. Koczkowska, M., Callens, T., Chen, Y., Gomes, A., Hicks, A.D., Sharp, A., Johns, E., Uhas, K.A., Armstrong, L., Bosanko, K.A., et al. (2020). Clinical spectrum of individuals with pathogenic NF1 missense variants affecting p.Met1149, p.Arg1276, and p.Lys1423: genotype-phenotype study in neurofibromatosis type 1. Hum Mutat 41, 299–315. 10.1002/humu.23929.

7. Evans, D.G., Baser, M.E., McGaughran, J., Sharif, S., Howard, E., and Moran, A. (2002). Malignant peripheral nerve sheath tumours in neurofibromatosis 1. J Med Genet 39, 311–314. 10.1136/jmg.39.5.311.

8. Uusitalo, E., Rantanen, M., Kallionpaa, R.A., Poyhonen, M., Leppavirta, J., Yla-Outinen, H., Riccardi, V.M., Pukkala, E., Pitkaniemi, J., Peltonen, S., and Peltonen, J. (2016). Distinctive Cancer Associations in Patients With Neurofibromatosis Type 1. J Clin Oncol 34, 1978–1986. 10.1200/JCO.2015.65.3576.

9. Gutmann, D.H., Rasmussen, S.A., Wolkenstein, P., MacCollin, M.M., Guha, A., Inskip, P.D., North, K.N., Poyhonen, M., Birch, P.H., and Friedman, J.M. (2002). Gliomas presenting after age 10 in individuals with neurofibromatosis type 1 (NF1). Neurology 59, 759–761. 10.1212/wnl.59.5.759.

10. Ferner, R.E. (2007). Neurofibromatosis 1 and neurofibromatosis 2: a twenty first century perspective. Lancet Neurol 6, 340–351. 10.1016/S1474-4422(07)70075-3.

11. Easton, D.F., Ponder, M.A., Huson, S.M., and Ponder, B.A. (1993). An analysis of variation in expression of neurofibromatosis (NF) type 1 (NF1): evidence for modifying genes. Am J Hum Genet 53, 305–313.

12. De Raedt, T., Brems, H., Wolkenstein, P., Vidaud, D., Pilotti, S., Perrone, F., Mautner, V., Frahm, S., Sciot, R., and Legius, E. (2003). Elevated risk for MPNST in NF1 microdeletion patients. Am J Hum Genet 72, 1288–1292. 10.1086/374821.

13. De Raedt, T., Brems, H., Lopez-Correa, C., Vermeesch, J.R., Marynen, P., and Legius, E. (2004). Genomic organization and evolution of the NF1 microdeletion region. Genomics 84, 346–360. 10.1016/j.ygeno.2004.03.006.

14. De Raedt, T., Beert, E., Pasmant, E., Luscan, A., Brems, H., Ortonne, N., Helin, K., Hornick, J.L., Mautner, V., Kehrer-Sawatzki, H., et al. (2014). PRC2 loss amplifies Ras-driven transcription and confers sensitivity to BRD4-based therapies. Nature 514, 247–251. 10.1038/nature13561.

15. Neefjes, J., Jongsma, M.L., Paul, P., and Bakke, O. (2011). Towards a systems understanding of MHC class I and MHC class II antigen presentation. Nat Rev Immunol 11, 823–836. 10.1038/nri3084.

16. Pagliuca, S., Gurnari, C., Rubio, M.T., Visconte, V., and Lenz, T.L. (2022). Individual HLA heterogeneity and its implications for cellular immune evasion in cancer and beyond. Front Immunol 13, 944872. 10.3389/fimmu.2022.944872.

17. Brosius, S.N., Sussman, J.H., DiStefano, I.A., Grothusen, G.P., Natan, K.N., Seka, I., Foltz, S., Labella, K., Bosse, K.R., Tan, K., and De Raedt, T. (2026). Identification of CD74-positive antigen presenting glioma cells in primary human tumors and murine models of NF1 high-grade glioma. Mol Cancer Ther. 10.1158/1535-7163.MCT-25-1215.

18. Orenbuch, R., Filip, I., Comito, D., Shaman, J., Pe’er, I., and Rabadan, R. (2020). arcasHLA: highresolution HLA typing from RNAseq. Bioinformatics 36, 33–40. 10.1093/bioinformatics/btz474.

19. Kochat, V., Raman, A.T., Landers, S.M., Tang, M., Schulz, J., Terranova, C., Landry, J.P., Bhalla, A.D., Beird, H.C., Wu, C.C., et al. (2021). Enhancer reprogramming in PRC2-deficient malignant peripheral nerve sheath tumors induces a targetable de-differentiated state. Acta Neuropathol 142, 565–590. 10.1007/s00401-021-02341-z.

20. Cortes-Ciriano, I., Steele, C.D., Piculell, K., Al-Ibraheemi, A., Eulo, V., Bui, M.M., Chatzipli, A., Dickson, B.C., Borcherding, D.C., Feber, A., et al. (2023). Genomic Patterns of Malignant Peripheral Nerve Sheath Tumor (MPNST) Evolution Correlate with Clinical Outcome and Are Detectable in Cell-Free DNA. Cancer Discov 13, 654–671. 10.1158/2159-8290.CD-22-0786.

21. Larsson, A.T., Bhatia, H., Calizo, A., Pollard, K., Zhang, X., Conniff, E., Tibbitts, J.F., Rono, E., Cummins, K., Osum, S.H., et al. (2023). Ex vivo to in vivo model of malignant peripheral nerve sheath tumors for precision oncology. Neuro Oncol 25, 2044–2057. 10.1093/neuonc/noad097.

22. Yan, J., Chen, Y., Patel, A.J., Warda, S., Lee, C.J., Nixon, B.G., Wong, E.W., Miranda-Roman, M.A., Yang, N., Wang, Y., et al. (2022). Tumor-intrinsic PRC2 inactivation drives a context-dependent immune-desert microenvironment and is sensitized by immunogenic viruses. J Clin Invest 132. 10.1172/JCI153437.

23. D’Angelo, F., Ceccarelli, M., Tala Garofano, L., Zhang, J., Frattini, V., Caruso, F.P., Lewis, G., Alfaro, K.D., Bauchet, L., et al. (2019). The molecular landscape of glioma in patients with Neurofibromatosis 1. Nat Med 25, 176–187. 10.1038/s41591-018-0263-8.

24. Fisher, M.J., Jones, D.T.W., Li, Y., Guo, X., Sonawane, P.S., Waanders, A.J., Phillips, J.J., Weiss, W.A., Resnick, A.C., Gosline, S., et al. (2021). Integrated molecular and clinical analysis of low-grade gliomas in children with neurofibromatosis type 1 (NF1). Acta Neuropathol 141, 605–617. 10.1007/s00401-021-02276-5.

25. Pan, Y., Hysinger, J.D., Barron, T., Schindler, N.F., Cobb, O., Guo, X., Yalcin, B., Anastasaki, C., Mulinyawe, S.B., Ponnuswami, A., et al. (2021). NF1 mutation drives neuronal activity-dependent initiation of optic glioma. Nature 594, 277–282. 10.1038/s41586-021-03580-6.

26. Geng, Z., Wafula, E., Corbett, R.J., Zhang, Y., Jin, R., Gaonkar, K.S., Shukla, S., Rathi, K.S., Hill, D., Lahiri, A., et al. (2025). The Open Pediatric Cancer Project. Gigascience 14. 10.1093/gigascience/giaf093.

27. Miller, D.T., Cortes-Ciriano, I., Pillay, N., Hirbe, A.C., Snuderl, M., Bui, M.M., Piculell, K., Al-Ibraheemi, A., Dickson, B.C., Hart, J., et al. (2020). Genomics of MPNST (GeM) Consortium: Rationale and Study Design for Multi-Omic Characterization of NF1-Associated and Sporadic MPNSTs. Genes (Basel) 11. 10.3390/genes11040387.

28. Mosaad, Y.M., Farag, R.E., Arafa, M.M., Eletreby, S., El-Alfy, H.A., Eldeek, B.S., and Tawhid, Z.M. (2010). Association of human leucocyte antigen Class I (HLA-A and HLA-B) with chronic hepatitis C virus infection in Egyptian patients. Scand J Immunol 72, 548–553. 10.1111/j.1365-3083.2010.02468.x.

29. Khor, S.S., Omae, Y., Nishida, N., Sugiyama, M., Kinoshita, N., Suzuki, T., Suzuki, M., Suzuki, S., Izumi, S., Hojo, M., et al. (2021). HLA-A*11:01:01:01, HLA-C*12:02:02:01-HLA-B*52:01:02:02, Age and Sex Are Associated With Severity of Japanese COVID-19 With Respiratory Failure. Front Immunol 12, 658570. 10.3389/fimmu.2021.658570.

30. Lorente, L., Martin, M.M., Franco, A., Barrios, Y., Caceres, J.J., Sole-Violan, J., Perez, A., Marcos, Y.R.J.A., Ramos-Gomez, L., Ojeda, N., et al. (2021). HLA genetic polymorphisms and prognosis of patients with COVID-19. Med Intensiva 45, 96–103. 10.1016/j.medin.2020.08.004.

31. Adebamowo, S.N., Adeyemo, A., Adebayo, A., Achara, P., Alabi, B., Bakare, R.A., Famooto, A.O., Obende, K., Offiong, R., Olaniyan, O., et al. (2024). Genome, HLA and polygenic risk score analyses for prevalent and persistent cervical human papillomavirus (HPV) infections. Eur J Hum Genet 32, 708–716. 10.1038/s41431-023-01521-7.

32. Villodre, E.S., Kipper, F.C., Pereira, M.B., and Lenz, G. (2016). Roles of OCT4 in tumorigenesis, cancer therapy resistance and prognosis. Cancer Treat Rev 51, 1–9. 10.1016/j.ctrv.2016.10.003.

33. Gangoso, E., Southgate, B., Bradley, L., Rus, S., Galvez-Cancino, F., McGivern, N., Guc, E., Kapourani, C.A., Byron, A., Ferguson, K.M., et al. (2021). Glioblastomas acquire myeloid-affiliated transcriptional programs via epigenetic immunoediting to elicit immune evasion. Cell 184, 2454–2470 e2426. 10.1016/j.cell.2021.03.023.

34. Johnson, A.M., Bullock, B.L., Neuwelt, A.J., Poczobutt, J.M., Kaspar, R.E., Li, H.Y., Kwak, J.W., Hopp, K., Weiser-Evans, M.C.M., Heasley, L.E., et al. (2020). Cancer Cell-Intrinsic Expression of MHC Class II Regulates the Immune Microenvironment and Response to Anti-PD-1 Therapy in Lung Adenocarcinoma. J Immunol 204, 2295–2307. 10.4049/jimmunol.1900778.

35. Chen, Y.Y., Chang, W.A., Lin, E.S., Chen, Y.J., and Kuo, P.L. (2019). Expressions of HLA Class II Genes in Cutaneous Melanoma Were Associated with Clinical Outcome: Bioinformatics Approaches and Systematic Analysis of Public Microarray and RNA-Seq Datasets. Diagnostics (Basel) 9. 10.3390/diagnostics9020059.

